# Diagnostic accuracy of a remote eye exam in measuring refractive errors and detecting cataract, glaucoma, and diabetic retinopathy

**DOI:** 10.1101/2023.11.09.23297101

**Authors:** Taís de Campos Moreira, Cassia Garcia Moraes Pagano, Maria Eulália Vinadé Chagas, Maicon Falavigna, Dimitris Rucks Varvaki Rados, Rodolfo Souza da Silva, Erno Harzheim, Marcelo Rodrigues Gonçalves, Roberto Nunes Umpierre, Pietro Baptista de Azevedo, Alice Paul Waquil, Caroline Fabris, Martha Pereira Lima Lang, Adriana Szortika, Anelise Szortyka, Aline Lutz de Araujo, Felipe Cezar Cabral

## Abstract

**Porpuse:** Approximately 2.2 billion people worldwide have some type of visual impairment and among these, many could have been avoided or solved if adequate care had been provided in a timely manner. In these cases, telemedicine can assist in solving the problems of many ophthalmological conditions by providing support, performing screening, telediagnosis or supervision of distant appointments. The use of telemedicine for eye health is well established for some eye diseases, such as diabetic retinopathy, macular diseases and strabismus, however, there is a lack of information on diagnostic accuracy for other ophthalmological pathologies prevalent in the world population, such as refractive errors. To evaluate the reliability of the diagnosis made through teleophthalmology, using the ophthalmologist’s face-to-face diagnosis as the gold standard.

**Methods:** A diagnostic accuracy study was carried out, with 216 patients assessed by the TeleOftalmo project who underwent eye examinations both on-site and through telemedicine. The patients underwent refraction tests, intraocular pressure measurement, photodocumentation in the slit-lamp and subjective refraction using the visual acuity screen and the digital refractor.

**Results:** The conditions evaluated were refractive errors, cataracts, diabetic retinopathy and suspected glaucoma. Kappa values, sensitivity, specificity, positive predictive value and negative predictive value were calculated. Approximately, 61.6% of the sample were women, with mean age of 47 years old. Kappa values show strong reliability regarding the diagnosis of cataracts (k = 0.86) and diabetic retinopathy (k = 0.79). Regarding refractive errors and suspected glaucoma, the kappa values were considered moderate (k = 0.598 and 0.478, respectively).

**Conclusion:** The use of telemedicine has shown favorable results regarding the diagnosis of refractive errors, cataracts and diabetic retinopathy, as well as moderate reliability for suspected glaucoma.

## Introduction

The telemedicine became a valuable tool during the SARS-CoV-2 pandemic, and its use has been more frequent since then^[1,2]^. Practitioners in different medical specialties started seeing patients online. In ophthalmology, the use of telemedicine reduces unnecessary outpatient visits^[3]^ and streamlines the care access of patients with eye conditions. For that purpose, in Brazil, telemedicine has shown to relieve one of the most pressing healthcare issues, which is the long waiting line for ophthalmology and other medical specialties. Now and even before the pandemic, telemedicine helps balancing the supply and demand for ophthalmology in underserved communities in a territory with physicians poorly distributed^[4]^.

Approximately 2.2 billion people worldwide have some type of visual impairment and among these, many could have been avoided or solved if adequate care had been provided in a timely manner^[5]^. Telemedicine can improve this scenario through different strategies, such as ocular health promotion initiatives, or by providing instructions in eye care to primary care physicians, or by virtually examining patients for screening, remote diagnosis, or remote follow-up visits ^[4,6]^.

One of the main concerns of doctors and patients with regards to teleophthalmology relates to the accuracy of a remote exam^[7]^.The use of telemedicine for eye health is well established for diabetic retinopathy and retinopathy of prematurity screenings ^[4,8–11]^. However, if eye care providers were to perform a comprehensive eye and vision assessment in a completely remote manner, there is not enough supporting evidence to date on how accurate such an evaluation would be. Thus, this study aimed to assess the reliability of diagnoses made through teleophthalmology, using the ophthalmologist’s in-person diagnosis (on-site assessment) as the gold standard. In this study, the same patients went through both evaluations, on-site and remote, consecutively. The outcomes of interest were suspected glaucoma, diabetic retinopathy, cataract, and refractive errors. For the latter, we compared the subjective refractions performed on-site and remotely, in order to assess the accuracy of the glass prescription from a distance.

## Methods

We carried out a prospective diagnostic accuracy study, and this report follows the Standards for Reporting Diagnostic Accuracy studies (STARD)^[12]^. The study protocol was approved by the Institutional Review Board of Hospital de Clínicas de Porto Alegre, in Porto Alegre – Rio Grande do Sul, Brazil.

### Study period

We prospectively enrolled patients and collected data over 13 weeks. The collection started in January 2020, was subsequently paused from March to August due to COVID-19 pandemic, and resumed aftwards.

### Settings

This study was conducted at TeleOftalmo, a telemedicine research and healthcare delivery program launched in 2017 in southern Brazil. The infrastructure includes a command center, where the attending physicians work, and eight remote or satellite units, where the patients go for their remote eye examination. The ophthalmologist at the command center has real-time audio and video communication with the satellite units and oversees the data acquisition by the local nursing team. For this study, we enrolled patients visiting one of TeleOftalmo’s satellite units, in Porto Alegre – Rio Grande do Sul, Brazil.

### Study participants

Consecutive patients arriving for a visit at the remote unit were invited to participate in the study. These patients were adults or children older than 7 years old. The exclusion criteria were pregnancy, acute red eye or vision loss, and diseases that require immediate intervention. Patients with cognitive impairment that prevented proper data collection would be excluded as well^[3]^. Patients were enrolled in the study after the written informed consent was obtained. From the participants between 7 to 18 years written informed consent was obtained from the parent/guardian.

### Data collection

We set up the research forms on the REDCap® platform^[13]^ and instructed the research teams to enter the data in the appropriate fields. REDCap was also set to randomize patients into the order they would be examined - if they would go through the on-site or remote examination first. The study workflow was tested in a pilot study to validate the logistics and data collection forms.

Patients were enrolled in the study in a consecutive way. The research coordinator entered each patient on the platform and assigned their exam order according to the randomization. Patients assigned to go through the telemedicine evaluation first were subsequently assessed by the on-site physician and vice versa. If the patient had undergone pupil dilatation (tropicamide 1% eyedrops), he/she returned to complete the assessment in both study arms (Figure 1).

**Figure 1.**
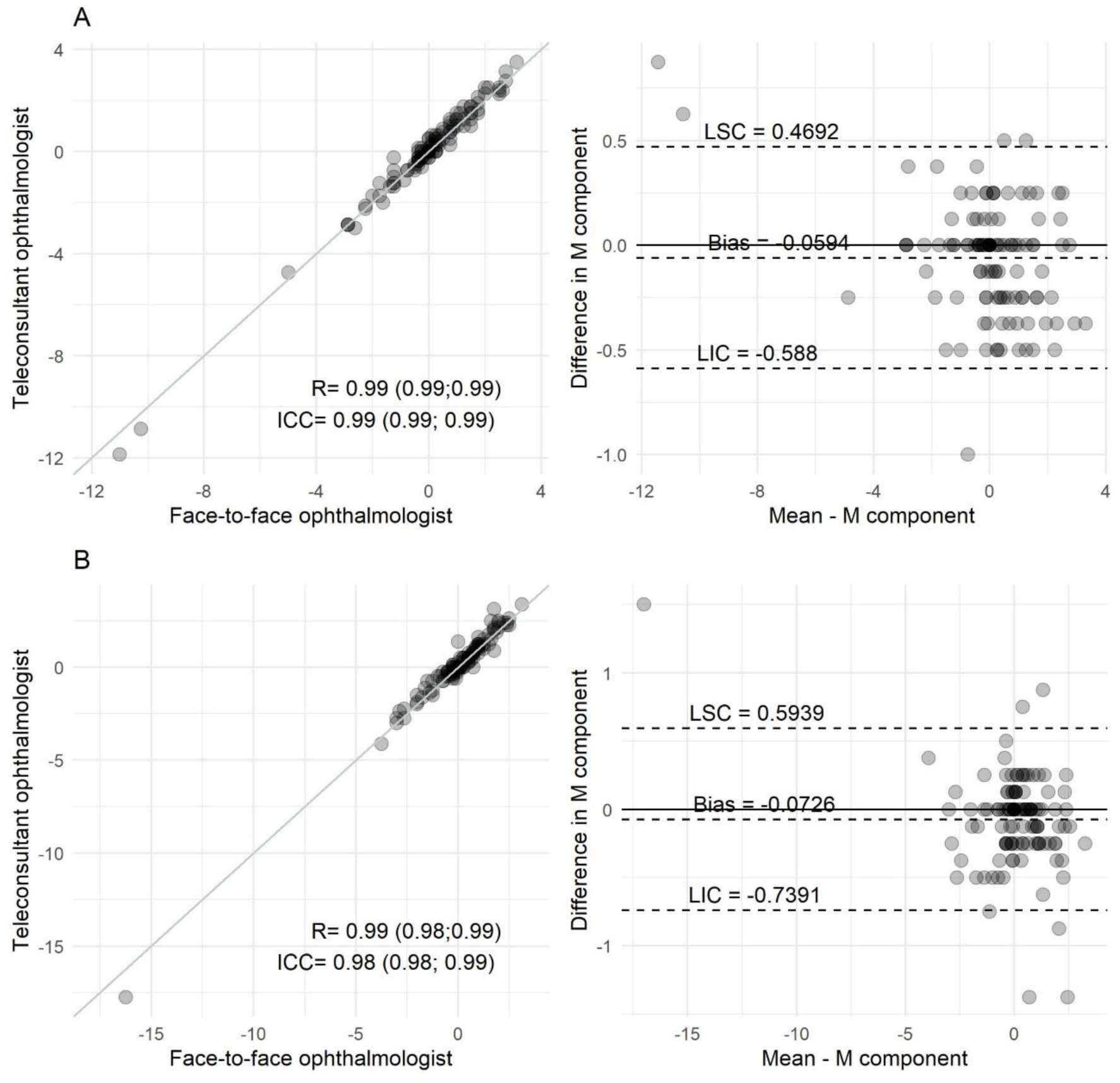
Correlation charts between the assessments and Bland-Altman comparing the assessments performed by ophthalmology physicians on-site and through telemedicine for: A. Spheric equivalent M for the right eye; B. Spheric equivalent M for the left eye.

#### 1. On-site assessment

The on-site assessment was carried out as a routine, comprehensive eye examination at the ophthalmologist office. It started with a brief clinical interview to understand the patient’s clinical history, such as eye diseases, medication use, family history of diseases, among others. Right after, the ophthalmologist assessed the following: visual acuity (VA), extraocular motility, pupillary reflexes, automated refractometry and keratometry (Visuref 100, Carl Zeiss®) air-puff tonometry (Visuplan 500, Carl Zeiss^®^), dynamic refraction with a digital refractor (Visuphor 500, Carl Zeiss^®^) biomicroscopy of the anterior segment, and ocular fundus examination with a 78D indirect ophthalmoscopy lens (SL 115, Carl Zeiss®).

#### 2. Telemedicine assessment

The telemedicine assessment started with the clinical interview by the nurse technician, who fills out the patient record on the telemedicine platform. The exams were taken following the usual procedures at TeleOftalmo: automated refractometry and keratometry (Visuref 100, Carl Zeiss^®^), air-puff tonometry (Visuplan 500, Carl Zeiss^®^), slit lamp (SL 115, Carl Zeiss^®^) photos of the eye’s anterior segment, and fundus images (Visucam 224, Carl Zeiss®). A detailed description of Teleoftalmo’s examination protocol is available elsewhere ^[14]^. In brief, the nurse technician takes 5 images of each patient’s eye, out of which 3 are on the slit lamp and 2 are on the retinal camera. The slit lamp photos consist of: (i) an 8x magnification view of the anterior chamber with direct, diffuse illumination to show a frontal view of the eye, (ii) a 12x-magnification view of the lens with direct, focal illumination, and (iii) a 12x-magnification view of the anterior chamber periphery with direct, focal illumination to assess the angle amplitude according to the Van Herick’s system^[15]^. The fundus photographs are one 45-degree centered at the macula, and one 30-degree centered at the optic disc. Photos were sent to the remote ophthalmologist using image sending software (Carl Zeiss®). The remote ophthalmologist synchronously reviewed the images and test results, and proceeded to the subjective refraction, which is performed with a digital VA screen and refractor that are operated from a distance. The ophthalmologist interacts with the patient through video-conferencing during the lens tests.

After the patient went through the on-site and telemedicine assessments, tropicamide eye drops 1% were instilled for peripheral retina assessment or static refraction, when requested by either physician. Upon completion of the assessments, the physicians took clinical notes and ticked the presence of each outcome on the RedCap platform. The outcomes of interest are defined in Table 1.

**Table 1.**
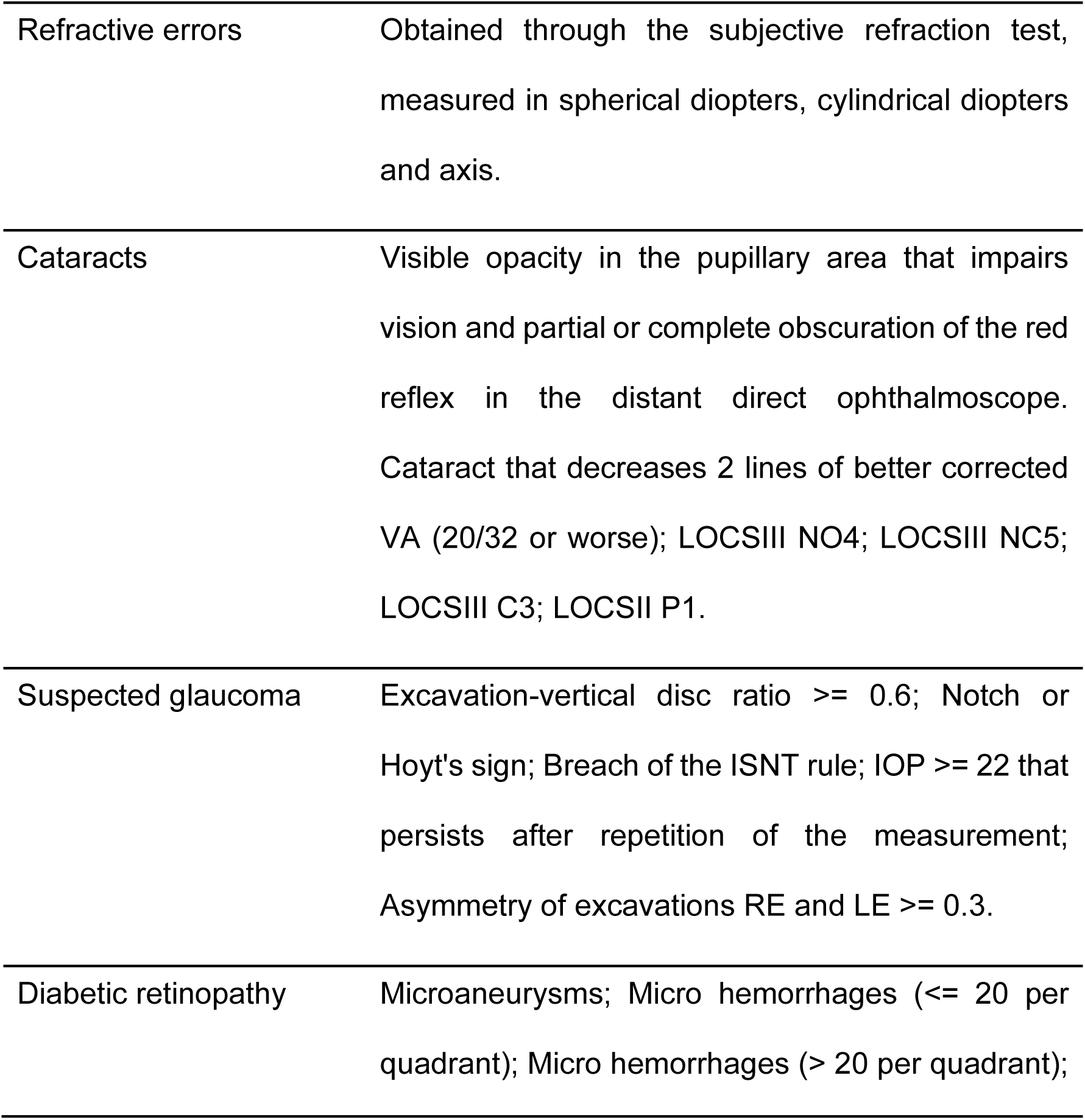
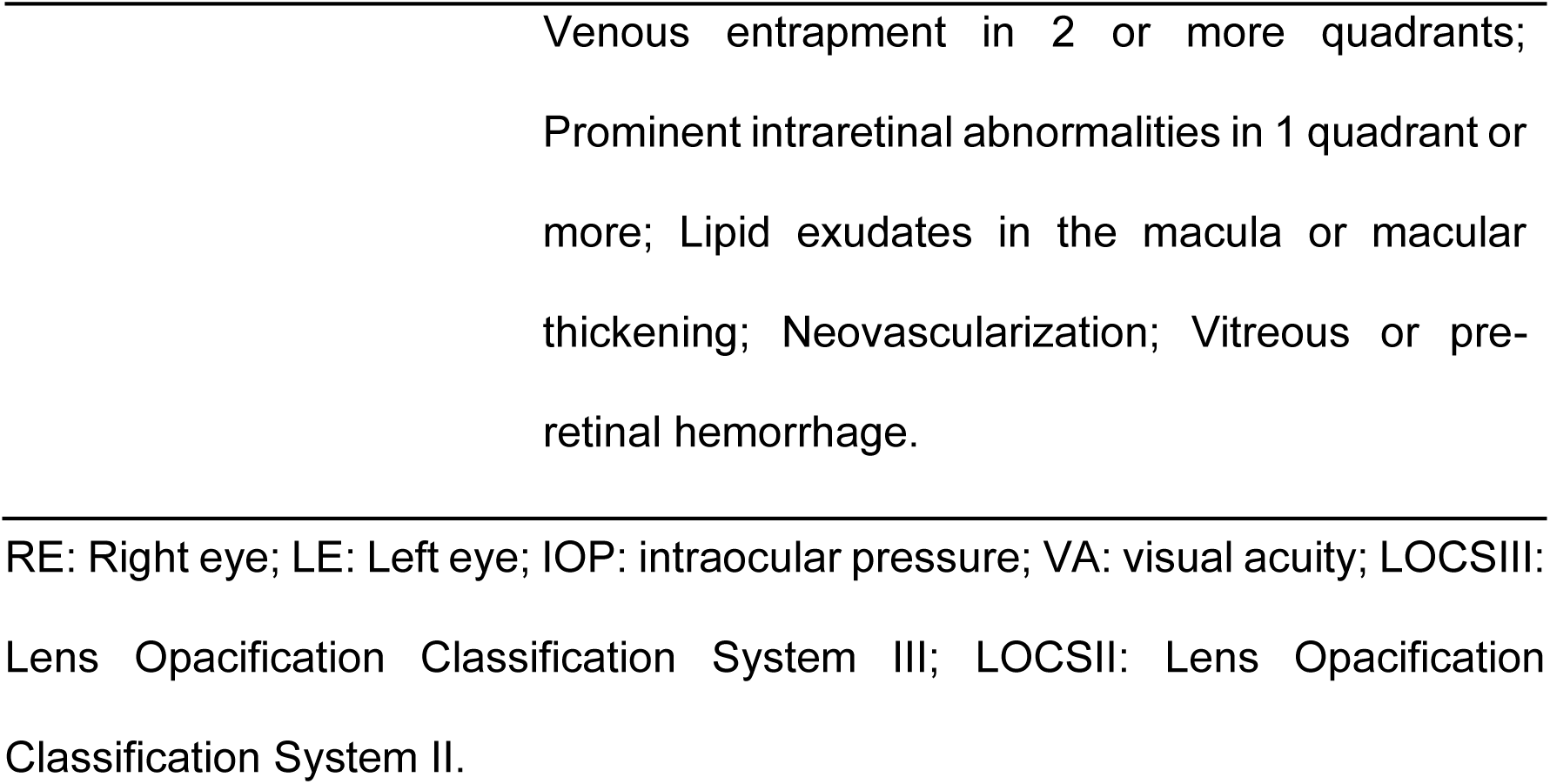
Definitions of ocular conditions.

Since both evaluations were carried out on the same day, some procedures were taken to minimize patients’ bench time while assuring that both physicians enjoyed similar access to the patients’ eyes and exams resources. These procedures include:

- Evaluation after pupildilation was performed in both arms between 20-30 minutes.
- Tonometry was taken only once for all patients. This was meant to avoid a possible confounding factor of repeated jet air onto the cornea, which can reduce further IOP measurements. If the attending physician (either on-site or remote) decided to repeat the IOP measurement, tonometry was taken after at least 15 minutes of the first measurement.
- Lensmeter determination was performed only once for all patients, for the sake of time.

### Sample size

All outcomes of interest were accounted in the sample size calculation. The largest sample size estimation was for suspected glaucoma, which was then adopted for this study.

The sample size was calculated assuming an estimated kappa agreement coefficient of 0.65^[16]^ for suspected glaucoma. The necessary sample size would then be 200 patients to achieve a significance level of 5%, considering accuracy of 15%^[17]^. Based on TeleOftalmo’s records, the expected prevalence of suspected glaucoma in the target population is 15%. Considering 5% losses, a sample size of 211 patients was necessary.

### Statistical analyses

Continuous variables were described as mean and standard deviation or median with interquartile range, while categorical variables were described as absolute and relative frequencies. Refraction components (sphere, cylinder, and axis) were converted into power vectors using the method described by Thibos *et al*., to obtain independent parameters M, J0, J45, and B in diopters (D)^[18]^. Bland-Altman and scatter plots were used to analyze the agreement between the on-site and the telemedicine assessments, as well as correlation and intraclass correlation analyses between the observers. To evaluate the ability of detecting cataract, retinopathy and suspected glaucoma from remote, we calculated the sensitivity, specificity, positive predictive value (PPV), negative predictive value (NPV), and interobserver agreement (Kappa) values, considering the ophthalmologist on-site assessment as the gold standard. In order to analyze refraction in a comparative way, the need of glass prescription was considered as an outcome, and then the remote performance for this outcome was calculated in the same wasy as the other outcomes. The R statistical package version 3.6.1^[19]^ was used to perform the analyses and the level of significance was set at 5%.

## Results

During the study period, 224 patients were enrolled in the study, out of which 8 were excluded. Thus, the final study population comprised 216 patients. The 8 exclusions were due to technical issues with the telemedicine system or because the patient left before the second assessment.

Complete demographics are described in Table 2. The majority (61.6%) were women, with a mean age of 47 years (SD ± 16.8).

**Table 2.** Demographic characteristics of the study population (n=216).

|  |  |  |
| --- | --- | --- |
| **Age, years** | Mean ± SD | 47.8 ± 16.8 |
|  | Range | 10.2 – 90.3 |
| n (%) | Teenagers | 20 (9.26) |
|  | Young adults | 10 (4.63) |
|  | Adults | 144 (66.67) |
|  | Older adults | 42 (19.44) |
| **Gender** | Female | 133 (61.57) |
|  | Male | 83 (38.43) |
| **Diabetes** | Yes | 19 (8.80) |
|  | No | 197 (91.2) |
| **Family history of glaucoma** | Yes | 24 (11.16) |
|  | No | 191 (88.94) |

The prevalence of eye diseases diagnosed by the on-site ophthalmologist was: cataracts (5.56%), suspected glaucoma (14.81%) and diabetic retinopathy (1.39%). The incidence of spectacle prescription was 77.31%, considering prescriptions to correct refractive errors of any kind. Comparisons among the on-site and the telemedicine assessments and diagnoses are described in Table 3. Noteworthy, there was disagreement on the need for glass prescription in 30 out of 216 cases, in which one arm prescribed new lenses and the other did not (16 on-site and 14 remote). This disagreement rate rendered a moderate kappa value (0.598), although the actual measurement of refraction was similar. Spherical equivalent (M) means differed between the on-site and the telemedicine assessment by -0.05 for the right eye (95% CI -0.10; -0.01, p-value = 0.017) and -0.07 for the left eye (95% CI -0.13; -0.01, p-value = 0.019). Bland-Altman plots for the right and left eye (fig 1) show the results for the spherical equivalent (M). The limits of agreement indicate that the differences range from -0.5 to 0.4 diopters for the right eye and from -0.7 to 0.5 for the left eye. The Bland-Altman graphs for cylindrical powers J0 and J45 can be found in the supporting information.

**Table 3.**
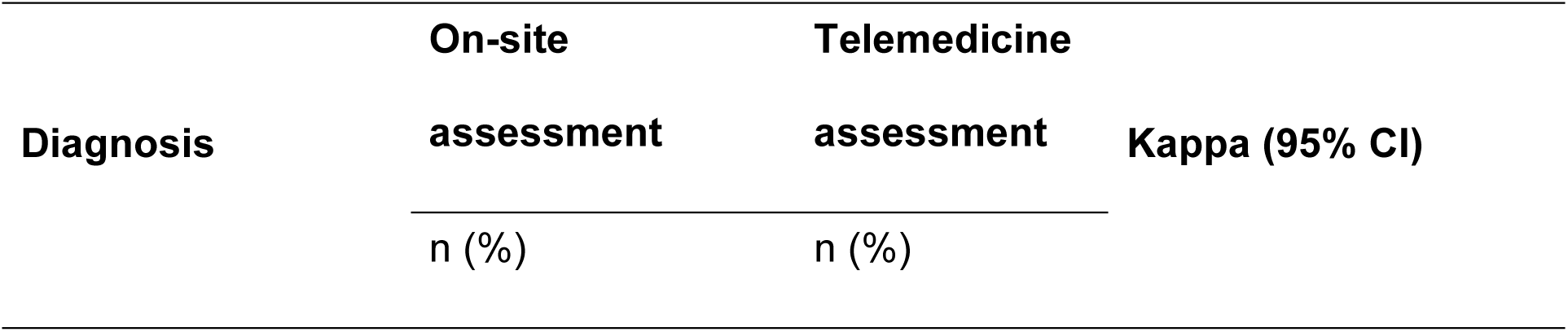
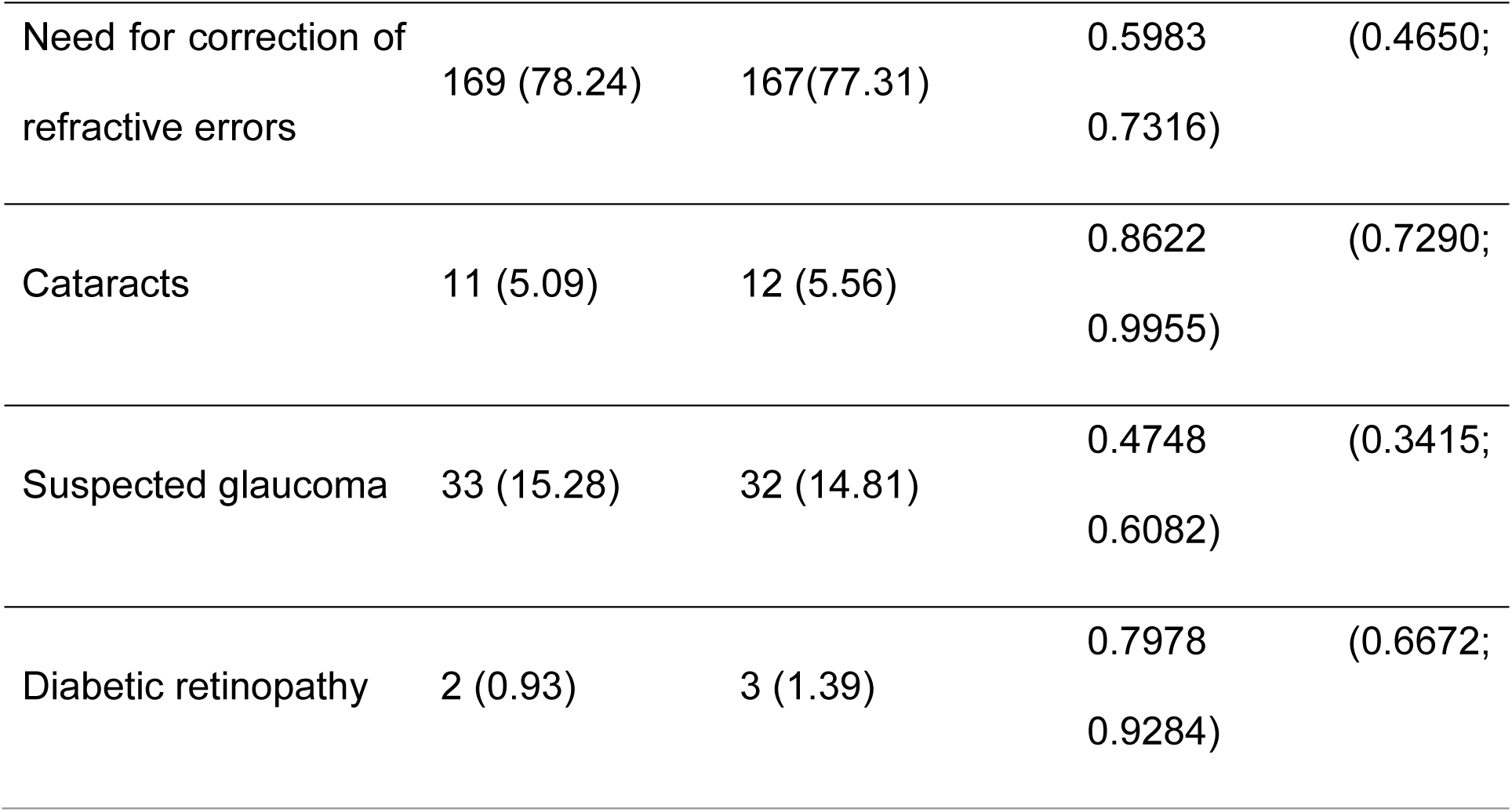
Prevalence of ophthalmologic conditions in the study population according to the diagnosis performed by an ophthalmologist on-site or through telemedicine and the agreement among the two diagnosis (n = 216).

Regarding the cataract detection, there was a strong diagnostic agreement between the on-site and the telemedicine assessments (kappa = 0.86). Concerning suspected glaucoma, the agreement was considered moderate (kappa = 0.478), due to a disagreement in29 cases (supporting information). For diabetic retinopathy the agreement was strong (kappa = 0.79)^[20]^ (Table 3).

The sensitivity of the teleophthalmology assessment to detect different clinical conditions ranged from 56% to 83%, depending on the condition. The positive predictive value was 55% to 100%. The specificity and negative predictive value were greater than 90% for all conditions. All accuracy values considered the entire cohort of 216 patients and are described in detail in Table 4.

**Table 4.** Sensitivity, specificity, positive predictive value, and negative predictive value of the ophthalmologist’s assessment through telemedicine compared to the assessment on-site (n = 216).

| **Diagnosis** | **Sensitivity**<br>**(95% CI)** | **Specificity**<br>**(95% CI)** | **PPV**<br>**(95% CI)** | **NPV**<br>**(95% CI)** |
| --- | --- | --- | --- | --- |
| Need for correction of refraction errors | 0.91<br>(0.85; 0.94) | 0.70<br>(0.55; 0.83) | 0.92<br>(0.86; 0.95) | 0.67<br>(0.52; 0.80) |
| Cataracts | 0.83<br>(0.52; 0.98) | 1.00<br>(0.97; 1.00) | 0.91<br>(0.59; 1.00) | 0.99<br>(0.97; 1.00) |
| Suspected glaucoma | 0.56<br>(0.38; 0.74) | 0.92<br>(0.87; 0.95) | 0.55<br>(0.36; 0.72) | 0.92<br>(0.87; 0.96) |
| Diabetic retinopathy | 0.67<br>(0.09; 0.99) | 1.00<br>(0.98; 1.00) | 1.00<br>(0.16; 1.00) | 1.00<br>(0.97; 1.00) |

NPV: negative predictive value; PPV: positive predictive value.

## Discussion

The results of this study are encouraging for the use of telemedicine in ophthalmology. We found a strong agreement between in-person and remote examinations for the detection of cataracts and diabetic retinopathy and a moderate agreement for suspected glaucoma. Refraction performed at a distance differed by less than 0.1 diopters in the spherical equivalent from the in-person test. This difference is very small and devoid of clinical significance. When accounting for the in-person findings as the gold standard, the sensitivity for the different findings ranged from 0.56 to 0.91 and the specificity from 0.70 to 1.00. The lowest sensitivity (i.e., 0.56) was found for the suspicion of glaucoma.

The outcomes of interest in this study represent the 4 leading causes of visual impairment in Brazil. Uncorrected refractive errors and cataracts are the most common ones, followed by diabetic retinopathy and glaucoma. Impaired access to specialty care has long been one of the greatest issues in the country’s public healthcare system, and telemedicine has started to play a role in alleviating the bottleneck for people in need of care.

Refractive errors alone affect nearly 60% of the population in Brazil, as found by Ferraz *et al*^[21]^. Our study demonstrates that the prescription made by a remote ophthalmologist with real-time tests is as accurate as the in-person ophthalmologist. One should note that glass prescription in Brazil is mainly performed by ophthalmologists and not optometrists. In this scenario, patients with refractive errors need a referral to an ophthalmology center just as patients with any other ocular condition. Telemedicine can increasingly assist in the detection and correction of refractive errors, especially in populations facing difficulties in accessing eye care^[22]^.

We compared subjective refraction values of the remote and on-site assessments. Refraction tests are affected by many factors such as the lens accommodation status and the patient’s mental state, including tiredness and anxiety. As such, small variations are expected, even if the exam is performed by the same doctor using the same equipment. Because of these variations, we chose Bland-Altman plots to compare the refraction components. One primary application of the Bland–Altman plot is to compare two clinical measurements -a new measurement technique against the gold standard, as even the gold standard is not without errors or deviations. The bias (or difference between the means) for the spherical equivalent was below 0.1, which is clinically irrelevant. To our knowledge, our study is the first to assess the accuracy of a real-time subjective refraction test performed at a distance. Other approaches use data collected by the patient themselves, or base the prescription on automatic refractometry. TeleOftalmo facilitates real-time virtual encounters, thus allowing the ophthalmologist to interact with the patient and perform subjective refraction tests.

There is robust evidence supporting the use of retinal fundus photographs to screen diabetic patients for retinopathy. Previous studies analyzed the accuracy of different readers, such as nurses, primary care physicians, and medical students. In our study, ophthalmologists that were not retina specialists evaluated the images, and the screening was part of a comprehensive eye exam that included visual acuity, slit lamp examination, and IOP measurement. We found a high accuracy for detecting diabetic retinopathy through telemedicine. However, the prevalence of diabetes in the study population was around 1%, because this study was designed for general ophthalmological visits and not specifically for diabetic patients. Previous publications showed an accuracy of around 90% or higher for the detection of diabetic retinopathy via teleophthalmology. Maa et al. in a similar teleophthalmological strategy found a positive predictive value of 100% and an agreement greater than 90% between remote read and on-site clinical examinations^[23]^. In a meta-analysis by Ullah, the overall sensitivity was 87% and the specificity was 91% for detecting diabetic retinopathy through retinal images^[24]^.

Sensitivity and specificity for suspected glaucoma was 86%. However, this should be interpreted with caution given the small number of positive cases in the sample. Remote ophthalmologists were more prone to suspect the presence of glaucoma than the in-person peers. Possible causes are that the remote ophthalmologist is not able to evaluate the optic disc in 3 dimensions, might have less confidence on the remote exam, or is following more strict criteria for referring patients. Previous literature recommends a high index of suspicion should be adopted for the diagnosis of glaucoma via telemedicine. Another study compared on-site assessments performed by a general ophthalmologist and telemedicine assessments performed by a glaucoma specialist. The sensitivity was 41.3% and the specificity of 95.8% for detecting glaucoma^[25]^. The level of agreement presented in these studies is relatively low, often because the early diagnosis of glaucoma is difficult and the criteria are rather subjective^[25,26]^. The outcome in our study was “glaucoma suspicion” instead of “glaucoma diagnosis”, because TeleOftalmo assessment did not include additional exams required to diagnose glaucoma (e.g., optical coherence tomography of the optic disc, computerized visual fields assessments). When it comes to glaucoma, telemedicine is more suitable for patients with initial cases and for monitoring disease evolution^[27]^, and a combination with facility visits is appropriate. In patients with more severe stages of glaucoma, on-site visits are still advised due to the risk of visual loss^[28]^. The main study limitation is the sample size. Several difficulties were faced in order to obtain a proper sample, the major one being the SARS-CoV-2 pandemic, which made it difficult to schedule patients’ visits, considering the need to follow safety rules to avoid contamination by the coronavirus (especially for people at increased risk of severe disease). Therefore, in order to properly clean the examination rooms, patients were scheduled with 30-minute intervals, slowing the data collection process. In addition, there was a small number of patients with some diagnoses, such as suspected glaucoma, increasing the size of the confidence intervals and causing the results to be interpreted with greater caution. Worthy of note, despite these limitations, many patients who underwent the telemedicine assessment had never been to an ophthalmology visit before or had been in the waiting line expecting for a visit for a long time.

Remote diagnosis in ophthalmology has the potential to play an important role in a broad population, as it is able to evaluate people that would otherwise face months-long waiting lines. Our findings show that teleophthalmology is possible and valid. The teleophthalmology strategy adopted by the TeleOftalmo project demonstrated accuracy results similar to the ones reported by other international experiences. It is important to highlight the need for training of professionals who are responsible for taking the ocular images and for their quality control, since the images have a central role in the practice of teleophthalmology^[14]^. The results described indicate that the TeleOftalmo project model, for comprehensive eye health care, can be replicated in other locations in Brazil, helping to reduce waiting lines and positively impacting the lives of patients, who are able to be diagnosed and receive proper care for potential ocular conditions.

## Data Availability

All data produced in the present work are contained in the manuscript

## Acknowledgements

To the nursing technicians Luana Ribeiro, Wagner da Rosa, Ana Paula Lopes, Daiana Michelli and Simone Previdi for assisting the patients and collecting data. To the nurse Marcio Gustavo Santanna da Silva for helping with data collection logistics and assisting the ophthalmology physicians. To the administrative technique Juliana Assis, Bárbara Carvalho De Oliveira e Veronica Guattini for helping with the schedules.

